# Discriminatory ability of adiposity phenotypes in identifying cardiometabolic disorders in indigenous and non-indigenous African populations

**DOI:** 10.1101/2024.09.07.24313232

**Authors:** Clement Nyuyki Kufe, Jean Claude Mbanya

## Abstract

**Background:** Whether any of the anthropometric indices are associated with cardiometabolic outcomes in indigenous Fulani African populations is not known. This study evaluates anthropometric indices in Fulani and non-autochthonous populations in predicting cardiometabolic outcomes in indigenous and non-indigenous populations.

**Methods:** A population-based cross-sectional study recruited 1 921 participants of similar median aged 32 (24–45) years from Fulbe (settled Fulani), Mbororo (nomadic pastoral Fulani) and the general population. Body weight (BW), height, waist circumference (WC), and hip circumference (HC) were measured and body mass index (BMI), waist-to-hip ratio (WHR), waist-to-height ratio (WHtR), Conicity Index (Cindex), body adiposity index (BAI), body roundness index (BRI) and body shape index (ABSI) were determined. The associations of anthropometric indices with cardiometabolic disorders were assessed by multivariable adjusted logistic regression and the area under the receiver-operating characteristic (ROC) curve compared the predictive abilities.

**Results:** Women had higher prevalence of MetS and the men had higher prevalence of hypertension and IFG/diabetes/hypertension while the prevalence of IFG/diabetes was similar in men and women. In women, the ROC and multivariable logistic regression analyses both showed consistent good performance of BW and BMI in identifying IFG/Diabetes in Fulbe and general population; BW, HC, BMI and BAI in Mbororo and BW, WC, HC, WHtR, and BRI in general population to predict hypertension and IFG/diabetes/hypertension. All the anthropometric indices showed good performance in all the groups to identify MetS. In men, WC, HC and BRI had good performance in all the groups while WC and BRI had good performance in all groups and HC in Fulbe and Mbororo to predict hypertension. The BW, WC, HC, BMI, WHtR, BAI and BRI showed consistent good performance in all the groups to predict MetS.

**Conclusion:** Anthropometric indices of adiposity are important risk assessing tools for cardiometabolic disorders in indigenous as well as non-autochthonous populations varying by ethnic group and sex.

## Introduction

Obesity and underweight might be symptomatic of an underlying health condition and malnutrition (1). Globally the prevalence of obesity (BMI≥30 kg/m^2^) increased from 3.2% to 10.8% in adult men and from 6.4 to 14.9% in adult women and underweight (BMI<18.5 kg/m^2^) reduced from 13.8 to 8.8% (2). This data does not include indigenous populations from Africa that have been shown to have high prevalence of underweight and low prevalence of obesity (3, 4). Typically weight and body mass index (BMI) have been used in clinical settings, though classifying risk based on body weight (BW) and BMI is insufficient due to large variability. Cardiometabolic risk prediction is useful in the identification of sub-phenotypes of metabolically unhealthy normal weight from metabolically healthy obesity using abdominal obesity indices. Increasing evidence have provided independent and additional information for inclusion of waist circumference (WC) in routine clinical practice (5). The BMI, WC and WHR has been shown not to improve cardiovascular risk prediction even with the inclusion of systolic blood pressure, history of diabetes and lipid levels in studies in developed countries (6). Other anthropometric such as the hip circumference (HC), waist-to-height ratio (WHtR), conicity index (Cindex) and body adiposity index (BAI) measurements are preferred for their simplicity and as such convenient to determine the cardiometabolic risk related to body adiposity. The BAI calculates the percentage of the body fat for adults of different ethnicities without using the (BW) (7). They do not require technical equipment and are used in resource limited settings. There is controversy over which anthropometric measurements best defines obesity and underweight and which best describes the risk gradient of cardiometabolic disturbance by sex and for each ethnic group.

Without population specification the use of these recommendations and cut off points is inapplicable. Epidemiologic studies suggest that these anthropometric measurements and criteria warrant a re-evaluation and appropriate ethnic specific modification proposed to suit diverse populations (8–11). These inconsistent findings warrant exploration of other anthropometric approaches that pose no risk to patients, and it remains unclear which sex-specific and ethnicity-specific anthropometric measurement has a superior discriminative power and should be adopted. Population specific cut-off points have been recommended but no assessment has been proposed for indigenous populations of Africa who generally lack health data. Previously we have shown that statistically significant differences exist between Mbororo and Fulbe for height, weight, WC, HC, BMI, BAI and between the general population and Fulbe for weight, WC, WHR, WHtR, BMI, CIndex, and fasting capillary glucose and the general population and Mbororo for weight, WC, HC, WHR, WHtR, BMI, Cindex, BAI and fasting capillary glucose and age and sex standardised prevalence of the cardiometabolic risk factors (3). There is a paucity of studies comparing anthropometric indices in different populations and by sex with the other common obesity markers.

Accordingly, this cross-sectional study aimed to identify the best anthropometric indicators for cardiometabolic disorders in the indigenous Fulani divided into settled Fulani known as Fulbe and nomadic pastoral Fulani known as Mbororo both sharing a common ancestry and language (Fulfulde), and the mixed non-indigenous population of the Bantu described as the general population. We compared the ability of the body weight, WC, HC, BMI, WHR, WHtR, Cindex, BAI, BRI and ABSI in discerning higher risk of IFG/diabetes, hypertension (HBP), IFG/diabetes/hypertension and metabolic syndrome (MetS).

### Study design and Methods

A cross-sectional study was conducted in five sites (Madjou II, Guiwa-Yangamou, Mazidou, Sabga and Gom-Mana) inhabited mostly by Fulani in the East and Adamawa regions of Cameroon. The detailed description of the study design and procedures has been described elsewhere (3,4). In summary, a multi-stage sampling procedure was used to recruit participants and a modified World Health Organisation (WHO) STEPwise approach questionnaire for the surveillance of NCDs was used to collect data. Data was collected on ethnic descent, behavioural risk factors included physical activity, diet and alcohol, tobacco consumption, health and medical history and family history of diabetes, hypertension and obesity. The body weight, height, waist and hip circumference were measured using standard procedures and Body Mass Index (BMI), Waist-Height Ratio (WHR), Waist-Height-Ratio (WHtR), Conicity Index (CIndex), Body Adiposity Index (BAI), BRI and ABSI were determined.

### Determination of anthropometric values

BMI was calculated as weight/height² (kg/m²) from weight and height (12). Central obesity was determined from the waist-to-hip-ratio (WHR) and waist-to-height ratio (WHtR) (13–15). The Conicity Index (Cindex) was calculated from waist circumference (m) ÷ 0.019 X √[Weight (Kg)/Height (m)] (16, 17). The Body Adiposity Index (BAI) (18) was obtained from (BAI = ((hip circumference in cm)/((height in meters) 1.5)-18)). The Body Roundness Index (BRI) combines height and WC to predict the percentage of regional and total fat was estimated from BRI=364.2 – 365.5 X √1 – ((WC/(2**π**))^2^/(0.5 x Height)^2^) (19) and body shape index reflecting body shape by incorporating WC, weight and height was obtained from ABSI= WC/(BMI^2/3^)X(Height^1/2^) (20).

### Blood pressure and fasting capillary glucose measurement

Blood pressure was measured on the right arm with uncrossed legs using arm blood pressure fully automated calibrated Omron M3 machine. Mean blood pressure of two closest measures was calculated. Fasting capillary glucose was measured with HemoCue Hb 201 DM Analyser (Angelhom, Sweden) (21) for participants who had no caloric intake for at least eight hours through an overnight fast.

### Definition of cardiometabolic disorders

#### IFG and Diabetes

International Diabetes Federation (IDF) and WHO diagnostic and classification criteria were used for two similar results with discordant results excluded for this analysis. Participants with fasting capillary glucose of 6.1–6.9mmol/L (110mg/dl–125mg/dl) were classified as impaired fasting glucose (IFG), ≥7mmol/L (126mg/dl) as diabetes and or on anti-diabetic medication (22).

#### Hypertension

Blood pressure for participants with mean Systolic Blood Pressure (SBP)≥130mmHg or mean Diastolic Blood Pressure (DBP)≥85mmHg or the two and/or self-reported current treatment of hypertension and/or taking anti-hypertensive medication were considered as hypertensive. Normal SBP was <120 mmHg and DBP <80 mmHg.

#### IFG/diabetes/hypertension

Participants with fasting capillary glucose of 6.1–6.9mmol/L (110mg/dl–125mg/dl) were classified as impaired fasting glucose (IFG), ≥7mmol/L (126mg/dl) as diabetes and or on anti-diabetic medication and Systolic Blood Pressure (SBP)≥130mmHg or mean Diastolic Blood Pressure (DBP)≥85mmHg or the two and/or self-reported current treatment of hypertension and/or taking anti-hypertensive medication were considered as hypertensive and classified as IFG/diabetes/hypertension.

#### Metabolic syndrome (MetS)

Metabolic syndrome was defined as elevated WC≥102 cm in men and ≥88cm in women, elevated blood pressure ≥130mmHg of SBP or ≥85mmHg of DBP or on antihypertensive drug treatment in a patient with a history of hypertension and elevated fasting capillary glucose of 6.1mmol/L (110mg/dl) or on drug treatment for elevated glucose (23).

### Data management and statistical analysis

Data was captured with Epi Data 3.0 and analysed using STATA 18.5 MP (StataCorp. College Station, TX: StataCorp LP). The Shapiro-Wilks test was used to assess the normal distribution of data. The socio-demographic, lifestyle factors, anthropometric measurements and metabolic disorder are described in the 3 population groups as frequencies and percentages for categorical data and median and 25^th^ to 75^th^ percentile for skewed continuous data as well as the mean for all normally distributed continuous variables as means and standard deviation. Spearman correlation coefficients (rho) were determined for each pair of the anthropometric measures and classified as very strong if rho was 0.8–1.0, strong if 0.6–0.8, moderate for 0.4–0.5, weak for >0.0–0.4 and no relationship if zero. Z-scores was calculated for each of the anthropometric values for the whole sample. The use of Z-scores permitted the comparison of the risk magnitude per 1 standard deviation (SD) change in the anthropometry, as well as sex-stratification using Fisher’s Yates transformation (24). The predictive ability of the anthropometric measurements for the cardiometabolic disorders were compared by the area under the curve (AUC) receiver-operating characteristic (ROC) curve and the 95% confidence interval (CIs) using the trapezoidal rule (25). The AUC of ≤0.5 reflects no discrimination, 0.6– 7 is considered moderate discrimination, 0.7–0.8 is acceptable and 0.8–0.9 excellent and >0.9 is outstanding (26, 27). Multiple logistic regression was used to evaluate the association between cardiometabolic disorder risk, and each standard deviation (SD) increase of the anthropometric measure adjusted for age group, marital status, physical activity, smoking and drinking status, fruit and vegetable consumption, salt and sugar intake and site and the multivariable adjusted odds ratios (MAORs) and 95% CI was reported. A p-value of <0.05 was considered statistically significant.

### Ethical approval

The study protocol was approved by National Ethics Committee of Cameroon, authorization No: 236/CNE/SE/2012. Informed consent was obtained from each participant by signing or thumb printing before inclusion in the study. Participants’ privacy and confidentiality during interview was guaranteed and data was anonymized and kept confidential. Statistical analysis was carried out on codified anonymous dataset. Each participant after counselling received confidential results of the anthropometric measures, blood pressure and fasting capillary glucose (FCG).

## Results

### Descriptive results

Data was analysed for 1 921 participants aged 32 (24–45) years, women 1 284 (66.8%) aged 30 (23–41) and men 637 (33.2%) aged 38 (27–50) years. The prevalence of body weight (BW) was significantly higher in the general population when compared to the Fulbe and Mbororo groups for men and women. The Mbororo women had the lowest BW [49 (43–56) kg, 53 (45–63) kg, 56 (50–65) kg, p=0.001] and BMI [(18.4 (16.7–20.8) kg/m^2^, 19.7 (17.3–22.9) kg/m^2^, 21.1 (19.3–24.0) kg/m^2^, p=0.001] when compared to the Fulbe and general population groups.

### Correlation between the anthropometric values by sex and group

The BW was strongly correlated with the WC, HC, BMI WHtR, BAI and BRI and weakly correlated with the Cindex and ABSI, p<0.05 in female and male of the Fulbe group except with the BAI in female that was moderate while the BW was strongly correlated with the WC, HC and BMI in the female and the BMI of the male in the Mbororo and BW was strongly correlated with the WC, HC and BMI of the female and males in the general population. The BW was not correlated with the WHR and Cindex of the female and male in the general population but weakly corrected in the female of the Mbororo population and weakly correlated with the male of the Mbororo but not with the Cindex. WC was strongly correlated with the HC, BMI, WHR, WHtR, Cindex and BRI in both the female and male and moderately correlated with ABSI in the female but not with ABSI of the Fulbe group while the WC was strongly correlated with the HC, BMI, WHR, WHtR, Cindex and BRI in the female of Mbororo and general population and with the WHR, WHtR, Cindex and BRI in the male of the Mbororo group and with the HC, BMI, WHtR, Cindex, and BRI of the male in general population group while the BAI and ABSI were moderately correlated with the WC in the Mbororo and the general population of both sexes. The HC was strongly correlated with BMI, WHtR, BAI and BRI, weakly correlated with Cindex and not correlated with WHR or ABSI in the female and male of the Fulbe group. The BMI was strongly correlated with the WHtR, BAI and BRI and weakly correlated with the WHR, and Cindex and not with the ABSI in the female and male of the Fulbe group. The WHR was strongly correlated with the WHtR, Cindex, BRI and ABSI and not with the BAI in both the male and female of the Fulbe group. The WHtR was strongly correlated with Cindex, BAI and BRI in the female and male but weakly correlated with the ABSI female and moderately in male. The Cindex was strongly correlated with BRI and ABSI and weakly correlated with the BAI in both female and male. The BAI was strongly correlated with BRI in both the female and male but weakly correlated with the ABSI in the female and not with the male in the Fulbe population while the ABSI was weakly correlated with the BRI in female and moderately correlated with the BRI in the male population of the Fulbe.

### Comparison of obesity phenotypes

The BW of women and men differed significantly for the Fulbe, Mbororo and general population (all, p<0.001). The WC and BMI of women and men was similar for the Fulbe (p>0.05), and general population (p>0.05) but was lower for the Mbororo women compared to the men (WC: 72.0 vs. 75.0, p=0.001 and BMI: 18.3 vs. 18.8, p=0.028). There was no difference in the HC of women and men for the Fulbe (p=0.987), and Mbororo (p=0.641) but women of the general population had as higher HC compared to the men (92.0 vs. 89.0, p=0.002). Women had lower WHR than men of the Fulbe (0.82 vs. 0.85, p=0.001) and Mbororo (0.82 vs. 0.86, p<0.001) but similar in the general population (p=0.233). The WHtR

### Comparison of the prevalence of cardiometabolic disorder (Figure 1)

The prevalence of IFG/diabetes was similar for women and men in Fulbe (15.0% vs. 16.6%, p=0.681), Mbororo (15.2% vs. 14.7%, p=0.843) and general population (15.6% vs. 17.5%, p=0.558). Women had significantly lower prevalence of hypertension than men, Fulbe (41.8% vs. 58.3%, p=0.001), Mbororo (40.2% vs. 53.2%, p<0.001), and the general population (40.9% vs. 50.5%, p=0.025). The prevalence of IFG/diabetes/hypertension was significantly lower in women than men, Fulbe (47.5% vs. 65.5%, p=0.001), Mbororo (49.4% vs. 59.4%, p=0.005) and general population (49.2% vs. 59.4%, p=0.017). Women had significantly higher prevalence of MetS than men, Fulbe (45.4% vs. 25.9%, p<0.001), Mbororo (34.8% vs. 21.0%, p<0.001) and the general population (52.7% vs. 26.9%, p<0.001).

**Figure 1:**
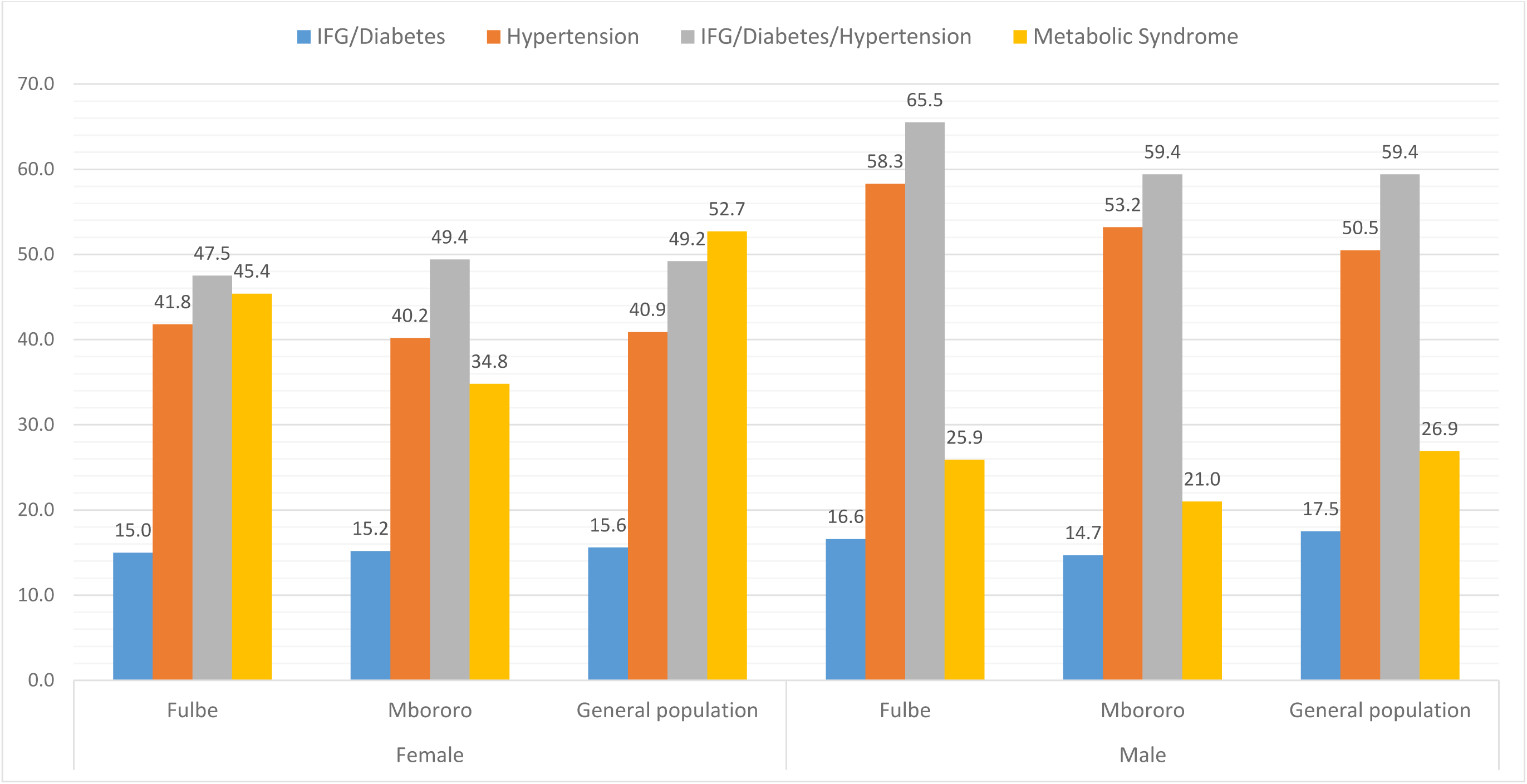
Prevalence of cardiometabolic disorder by group and sex.

There was no significant difference in the prevalence of IFG/diabetes for the three groups for women (p=0.976) and men (p=0.694); the prevalence of hypertension for women (p=0.901) and for men (p=0.356) as well as for IFG/diabetes/hypertension for women (p=0.866) and men (p=0.437). There were significant differences in the prevalence of MetS in the three groups for women (Fulbe: 45.4%, Mbororo; 34.8% & general population; 52.7%, p<0.001) and not for men (Fulbe; 25.9%, Mbororo; 21.0% & general population; 26.9%, p=0.263).

### Predictors of IFG/Diabetes

The area under the ROC curve showed moderate discrimination for the BW, WC, HC, BMI, WHR, WHtR, BAI and BRI in women and all the anthropometric measures in men of the Fulbe group. For the Mbororo group, the BMI and BAI in women while in men; the BW, WC WHR WHtR, Cindex and BRI showed moderate discrimination. The BMI, WHR, WHtR and BRI in women and WC, WHR, WHtR, Cindex, BRI and ABSI of the general population showed moderate discrimination. The area under the curve ROC analysis consistently showed moderate AUC values and highest MAORs for BW and BMI, all of which were statistically significant as the best predictor in identifying IFG/Diabetes in women of the Fulbe group and general populations and none for men. The WC, HC, WHR, WHtR, Cindex, BAI, BRI and ABSI were the worse predictors of IFG/diabetes in both women and men of the Fulbe, Mbororo and general population.

### Predictors of hypertension

From the area under the ROC curve, the BW, WC and HC showed moderate discrimination in women while BW, BMI WHtR and Cindex showed moderate discrimination and WC, HC, BAI and BRI showed acceptable discrimination in men of the Fulbe group in identifying hypertension. ROC analysis showed moderate discrimination for BW, WC, HC, BMI, WHtR, BAI and BRI had moderate discrimination for women while BW, WC, HC, BMI, WHR, WHtR, Cindex, BRI and ABSI in men of the Mbororo group. For the general population, all the anthropometric measures showed moderate discrimination in women and men except for ABSI in men. ROC curves consistently showed moderate AUC values and highest MAORs for BW, HC, BMI, and BAI in women and in men WC, HC and BRI in the Mbororo group. For the general population, the ROC analysis consistently showed best predicting capacity with BW, WC, HC, WHtR, Cindex, BAI and BRI in women and with BW, WC, HC, BMI, WHtR, BAI and BRI in men. However, none of the anthropometric measures identified hypertension in the women but BW, WC, HC, BMI, WHR, BAI and BRI consistently showed best predicting capacity with ROC analysis and MAORs in men of the Fulbe group. The ABSI was the worse predictor of hypertension in both women and men for all three groups.

### Predictors of IFG/diabetes/hypertension

For the Fulbe group, the area under the ROC curve did not show any discrimination characteristics as well as MAORs for all the anthropometric measures in women but showed moderate discrimination for WHR and Cindex and acceptable discrimination for BW, WC, HC, BMI, WHtR, BAI, and BRI in men. The ROC analysis showed moderate discrimination for BW, WC, HC, BMI, WHtR, BAI and BRI in women as well as all the anthropometric measures except ABSI in men of the Mbororo group. In the general population group the ROC analysis had moderate discrimination for all the anthropometric measures in men and women except ABSI in women. The BW, HC, BMI, BAI in women and WC, HC and BRI consistently had best predictor in men of the Mbororo group while BW, WC, HC, BMI, WHtR, BAI and BRI were best predictors in only men of the Fulbe group. While BW, WC, HC, BMI, WHtR and BRI were the best predictors in women, the WC, BMI, WHtR, BAI and BRI were best predictors in men of the general population. The WHR and ABSI were the worse predictors of IFG/diabetes/hypertension in both women and men of the Fulbe, Mbororo and general population.

### Predictors of MetS

The area under the ROC curve showed acceptable discrimination for BW, HC, BMI, WHR, Cindex, BAI, ABSI, excellent discrimination for WC, WHtR and BRI in the women and acceptable discrimination for BW, WC, HC, BMI, WHR, WHtR, Cindex, BAI, BRI, and ABSI in men of the Fulbe. For the Mbororo, ROC analysis had acceptable discrimination for BW, HC, BMI, WHR, Cindex, BAI, and ABSI and excellent discrimination for WC, WHtR and BRI in women and in men, moderate discrimination in all the anthropometric measures. For the general population, the ABSI showed moderate discriminatory while BW, HC, BMI, WHR, Cindex and BAI had acceptable discrimination and WC, WHtR and BRI had excellent discrimination in women while in men the BW, WHR, Cindex, and ABSI showed moderate discrimination, the WC, HC, BMI, WHtR, BAI and BRI had acceptable discrimination, and no excellent discrimination was showed for MetS in men.

All the anthropometric measures consistently showed good performance in predicting MetS in women while in men all except ABSI consistently showed good performance in predicting MetS in the Fulbe and apart from WHR, Cindex and ABSI, all showed good performance in predicting MetS in the Mbororo and the general population.

## Discussion

The key findings of the study were that all the anthropometric measures had good performance in predicting MetS risk in women while BW, WC, HC BMI and WHtR, BAI and BRI showed best performance in predicting MetS for the men in all the groups. The BW, HC and BMI had best performance in women of the Mbororo and general population and not Fulbe while WC and BRI showed best performance in all the three groups of men in predicting IFG/diabetes/hypertension. The BW, HC and BAI had best performance in women of the Mbororo and general population and not Fulbe while in men, the WC, HC and BRI showed best performance in predicting hypertension. Only the BW and BMI had best performance in women of Fulbe and general population in predicting IFG/diabetes. The worse performance in predicting IFG/diabetes, hypertension and IFG/diabetes/hypertension was from WHR and ABSI in both men and women of all the groups. WC and BRI are better predictors of hypertension and IFG/diabetes/hypertension in men in all the groups but not in women of all the groups.

While BW, WC, HC, BMI, WHtR, BAI and BRI were the best predictors of MetS in both men and women in the Fulbe, Mbororo and the general population. This is in line with the results of a study in Chinese men and women which identified anthropometric measures of abdominal obesity, especially WHtR in identifying MetS, IFG/diabetes and dyslipidemia and BMI for identifying people with hypertension and hyperuricemia in men (28) whereas our study suggested BMI for identifying women of the Mbororo group and men of the Fulbe and the general population at risk of hypertension. Using ROC analysis, WC and WHtR in our study for men and women of the Fulbe, Mbororo and general population and BMI in women of all the three groups and in men of the Fulbe predicted MetS. This was similar to a Chinese study that identified WC, BMI and WHtR to predict ≥3 metabolic risk factors in men and WC and BMI to predict ≥3 metabolic risk factors in women (29). Similar findings have been reported in a study of 202 Southern-Indians using area under the ROC curve analysis and adjusted odds ratios for age, sex, BMI, history of smoking and alcohol intake showed that BRI was superior in identifying MetS as well as Cindex if only area under the ROC curve analysis was considered with variations after stratification by sex (30). Similar findings were reported in a European study that analysed data of 12 328 participants, where using AUC analysis the BMI and WC in men and WHtR and WC in women was shown to have a better discriminatory power in identifying MetS except ABSI (31).

Our findings showed that WHtR was the best predictor for hypertension in women and men of the general population and women of the Fulbe group. Similar findings to our study were reported in the study of indigenous population of Orang Alsi villages in the Krau Wildlife Reserve, Pahang, Peninsular Malaysia where WHtR was the best predictor for hypertension (32). Contrary to the Orang Alsi indigenous populations we showed in that BW, HC and BAI in women of the Mbororo indigenous group and WC, HC and BRI in men of the Fulbe and Mbororo indigenous populations were equally good predictors of hypertension. This study showed similar findings to a study in Brazil for WC and WHtR as best predictors for hypertension in women of the general population (33). On the contrary our study also showed that WC, BMI and WHtR were also good predictors of hypertension in the men of the Fulbe and the general population.

Using area under the ROC curve analysis WC and WHtR were the best predictors of IFG (>110 mg/dL) in a study of 12 294 adults attending annual physical exams provided by EHE International in United States (34). This is in line with our ROC curve results of IFG/diabetes for women and men of the Fulbe group in our study. On further analysis of MAORs, neither WC nor WHtR was associated with the IFG/diabetes in our study.

Results of area under the ROC curve showed that measures of central adiposity such as WC, WHtR and BRI (measure of central adiposity) were consistently better predictors of MetS in both sexes than BMI (measure of overall adiposity) suggesting that complications of obesity are more closely related to body fat distribution rather than absolute degree of adiposity per se. This is consistent with other finding in a Singaporean study of Chinese, Malay and Indian adults suggesting that measures of central adiposity are associated CVD risk factors than BMI (35). Further analysis using MAORs investigating the effect of central adiposity on the overall adiposity showed that for each SD increase of WC, WHtR and BRI had significant ORs for MetS suggesting that measures of central adiposity increase the odds of cardiometabolic risk independent of BMI in women and men of all the groups and for hypertension and IFG/diabetes/hypertension of women in the general population and men of Fulbe, Mbororo and general population. We showed that WC, WHtR and BRI perform better in predicting MetS in both women and men of all the groups. BAI estimates percentage adiposity directly and similar to the Singaporean study, our study showed that it may be a good predictor of cardiometabolic disorders but not better than BMI (35).

A study of the Inuit populations in Nunavik (Northen Quebec, Canada) using AUC analysis reported an acceptable capacity of BW, BMI, WC and WHtR to predict hypertension in women and men similar to our study that showed a moderate acceptance in men of the Fulbe and Mbororo indigenous populations and women of the Mbororo indigenous populations (36). The difference may be explained by the differences in socio-economic status, nutrition and epidemiologic transition and genetics between the indigenous Fulani populations of Africa and the Inuit populations in Canada.

Some studies have suggested WHtR (37, 38), some prefer WC (39, 40) and WHR (41), others all of them (42), some a combination (43, 44) and another concluded that whether singly or in combination discriminating by sex (45), while some suggest no improvement in the prediction of cardiovascular disease risk (6). For the diverse African populations, the various optimal WC cut off for the identification of cardiometabolic risk by sex have been proposed from ≥78 cm to ≥80 cm for men and ≥82 cm to ≥85 cm for women and 96.8 cm for both dysglycaemia and type 2 diabetes for men (46) and 91.8 cm for incident dysglycaemia and 95.8 cm for type 2 diabetes for women (47) and ≥81.2 cm for men and ≥ 81.0 cm for women (48). WC often signify visceral adiposity is as important as the BMI if not more informative in individuals with elevated WC is associated with increased cardiometabolic risk in adults. Independent of sex and age, WC is associated with health outcomes within all BMI categories with the strength of the association between WC and morbidity and / or mortality after adjusting for BMI. When WC and BMI are used as continuous variables in risk prediction models, WC remains as a positive predictor of risk of death, but BMI is not or negatively related to the risk of death (49). To overcome the limitations of the traditional obesity markers, a body shape index (ABSI) combining the weight, height and WC and body roundness index (BRI) using height and WC to estimate the percentage of regional and total fat have been developed (50–53). Studies have shown that ABSI and BRI are associated with abdominal adipose tissue and cardiometabolic risk, onset of type 2 diabetes, premature mortality hazards than BMI and WC (28, 29, 54–55) and that BRI is even a better predictor of cardiometabolic risk in Southern-Indian adults and Eastern-China adults (30, 56). BRI and WHR have been shown to be a better predictor of CVD risk factors as well as WHtR and WC in assessing hypertension (57). A meta-analysis reported that BRI and ABSI have a discriminatory power for hypertension with BRI significantly a better predictor of hypertension than ABSI (58).

Upon stratification by sex, we observed variation in the correlation of the anthropometric measures, area under the ROC curve and MAORs analysis. The differences in BW, WC and HC and the general body fat distribution in women and men may explain the relative performance of the anthropometric measures. Further, the BW, WC, WC and height are direct measures used to calculate the BMI, WHR, WHtR, Cindex, BAI, BRI and ABSI. While Cindex is similar to the WHR in being a health indicator, the Cindex formula adjusts for WC for height and BW not requiring HC for body fat distribution (59). The use of HC in the BAI formula allowed for sex differences in adiposity to be considered when compared to BMI calculation that uses BW and height. BAI had good predictive power in the women of our study and Southern-Indian women and a lower predictive power in men (30). This may be explained by the lower adiposity in men as compared to women.

### Strengths and limitations of the study

Strengths of the study included a response rate of more than 90% for this quantitative population-based study. The study focused on an indigenous population. Risks factors are analysed for participants of indigenous Fulani populations from the settled Fulani (Fulbe) and the nomadic pastoral Fulani (Mbororo) and compared with the general population. The study involved risk factor analysis centred on individuals as a preponderant scale in probing epidemiological questions and adequately explaining who is at risk. The area under the ROC curve and multivariable adjusted logistic regression were used to assess the discriminatory capabilities. We standardised the estimates by the use of Z-scores which permitted us to compare the risk magnitude per 1 standard deviation (SD) change. Standardised WHO questionnaires and international guidelines were used for the definition of diabetes, hypertension, classes of overweight and obesity. The study provides baseline data on risk factors of indigenous populations. Trained enumerators conversant with the widely spoken local Fulfulde language administered the questionnaires. Anthropometric measurements such as body weight, height, WC and HC and calculations such as BMI are less likely to have measurement errors (60). However, evidence suggests measurements in body weight, WC and HC are prone to higher proportions of errors in obese populations (61) which was less likely in this study as the obesity levels were low. This is a cross-sectional study and limited to examining multiple scale causal mechanisms. Findings should be interpreted with caution and therefore does not allow for causal deductions. Accurate prediction of disease risk is difficult in specific population subtypes due to complexity of fundamental causes of diseases such as social, environmental drivers and genetic factors and interactions with environments and life course trajectories (62, 63).

### Conclusions

Anthropometric indices are important risk assessing tools for cardiometabolic disorders in indigenous as well as non-autochthonous populations. The findings show variation in the discriminatory capabilities of anthropometric measures by ethnicity and sex and the importance of using other indicators apart from the BMI which has the advantage of being simple, easy and cost-effective in resource limited settings. The BW, WC, HC, WHtR, BAI and BRI in addition to BMI are better in assessing MetS in indigenous as well as non-indigenous African populations and may not depend on sex. When stratified by sex, WC and BRI is a better alternative for detecting risk from hypertension and IFG/Diabetes/HBP in men of the indigenous Fulbe and Mbororo populations as well as in the women and men of the general population while the BW and BMI may be appropriate to predict IFG/Diabetes in women of the Fulbe and general populations. WHR, Cindex and ABSI were the worse anthropometric measures in predicting IFG/diabetes, hypertension and IFG/diabetes/hypertension in women and men of all groups while ABSI was the worse in detecting MetS in the men all the groups. With rising overweight and obesity rates, further research is needed to develop specific and culturally sensitive interventions in indigenous Fulani as well as non-indigenous populations to understand the complex interplay between adiposity and cardiometabolic disorders.

## Data Availability

All data produced in the present study are available upon reasonable request to the authors

## Competing interest

The authors have no competing interest to declare.

## Authors’ contributions

JCM and NCK reviewed the article and critically reviewed the article. NCK designed the analytic strategy, analysed the data and wrote the first and subsequent drafts. All authors contributed to the design, data collection and subsequent drafts which were read, reviewed and approved.

## Acknowledgments

We acknowledge input of HoPiT staff that participated in data collection and entry and especially research participants. We are grateful to local guides, village champions and chiefs and regional health representatives, district medical officers and local health staff

## Funding

Funding was provided by World Diabetes Foundation project number WDF12-707 to Health of Populations in Transition (HoPiT) Research Group, Cameroon.

## Role of the funding partner

The sponsor of the project did not play any role in the study design, data collection, analysis, interpretation and writing of the project report or this article. NCK had full access to all the data of the quantitative study, wrote and submitted the article for publication.

---

**Table 1:** Description of the 1 921 participants.

| **Sex, n (%)** | **Female, 1284 (66.8)** |  |  | **Male, 637 (33.2)** |  |  |
| --- | --- | --- | --- | --- | --- | --- |
| **Ethnicity, n (%)** | **Fulbe,**<br>**280 (21.8)** | **Mbororo, 632**<br>**(49.2)** | **General population,**<br>**372 (29.0)** | **Fulbe,**<br>**139 (21.8)** | **Mbororo,**<br>**286 (44.9)** | **General population,**<br>**212 (33.3)** |
| Age in years, median (25<sup>th</sup> – 75<sup>th</sup>) | 30 (23 - 40) | 30 (24 - 40) | 30 (23 - 43) | 37 (24 - 51) | 38 (28 - 52) | 35.5 (26 - 50) |
| **Age group, years, n (%)** |  |  |  |  |  |  |
| 20 – 39 | 197 (70.4) | 445 (70.4) | 255 (68.6) | 77 (55.4) | 151 (52.8) | 121 (57.1) |
| 40 - 59 | 67 (23.9) | 145 (22.9) | 84 (22.6) | 38 (27.3) | 98 (34.3) | 64 (30.2) |
| ≥ 60 | 16 (5.7) | 42 (6.7) | 33 (8.9) | 24 (17.3) | 37 (12.9) | 27 (12.7) |
| Smoker, n (%) | 5 (1.8) | 15 (2.4) | 25 (6.7) | 27 (19.4) | 49 (17.1) | 68 (32.1) |
| Ever drank alcohol, n (%) | 11 (3.9) | 21 (3.3) | 205 (55.1) | 24 (17.3) | 32 (11.2) | 132 (62.3) |
| **Fruit consumption** |  |  |  |  |  |  |
| High (5 – 7days/week) | 70 (25.0) | 101 (16.0) | 78 (21.0) | 34 (24.5) | 45 (15.7) | 49 (23.1) |
| Moderate (3 – 4 days/week) | 64 (22.9) | 149 (23.6) | 84 (22.6) | 39 (28.1) | 77 (26.9) | 54 (25.5) |
| Low (0 – 2 days/week) | 146 (52.1) | 382 (60.4) | 210 (56.5) | 66 (47.5) | 164 (57.3) | 109 (51.4) |
| **Vegetable consumption** |  |  |  |  |  |  |
| High (5 – 7days/week) | 178 (63.6) | 320 (50.6) | 162 (43.6) | 82 (59.0) | 150 (52.4) | 82 (38.7) |
| Moderate (3 – 4 days/week) | 77 (27.5) | 210 (33.2) | 133 (35.7) | 41 (29.5) | 91 (31.8) | 80 (37.7) |
| Low (0 – 2 days/week) | 25 (8.9) | 102 (16.1) | 77 (20.7) | 16 (11.5) | 45 (15.7) | 50 (23.6) |
| Always add salt to food | 54 (19.3) | 112 (17.7) | 87 (23.4) | 23 (16.6) | 50 (17.5) | 34 (16.0) |
| Always add sugar to tea/coffee | 51 (18.2) | 94 (14.9) | 94 (25.3) | 31 (22.3) | 58 (20.3) | 56 (26.4) |
| **10 mins Physical activity at work, n (%)** |  |  |  |  |  |  |
| Vigorous | 101 (36.1) | 235 (37.2) | 166 (44.6) | 63 (45.3) | 148 (51.8) | 135 (63.7) |
| Moderate | 79 (28.2) | 205 (32.4) | 130 (35.0) | 34 (24.5) | 66 (23.1) | 39 (18.4) |
| Low | 100 (35.7) | 192 (30.4) | 76 (20.4) | 42 (30.2) | 72 (25.2) | 38 (17.9) |
| **Anthropometry, median (25<sup>th</sup> – 75<sup>th</sup>)** |  |  |  |  |  |  |
| Body Weight (BW) | 50.0 (43.0 – 59.5) | 46.0 (41.0 – 53.0) | 54.0 (49.0 – 62.0) | 57.0 (52.0 – 68.0) | 54.0 (48.0 – 61.0) | 60.0 (54.0 – 67.0) |
| Waist circumference (WC) | 74.0 (66.5 – 84.0) | 72.0 (66.0 – 80.0) | 78.0 (72.0 – 85.5) | 76.0 (70.0 – 83.0) | 75.0 (70.0 – 81.0) | 76.0 (72.0 – 84.0) |
| Hip circumference (HC) | 90.0 (84.0 – 98.0) | 87.0 (83.0 – 93.0) | 92.0 (86.0 – 98.0) | 90.0 (86.0 – 97.0) | 88.0 (83.0 – 92.0) | 89.0 (85.0 – 96.0) |
| Body mass index (BMI) | 19.3 (16.9–23.0) | 18.3 (16.5–20.6) | 21.1 (19.4 – 24.5) | 20.1 (18.0 – 22.9) | 18.8 (17 – 21.1) | 21.2 (19.1 – 23.4) |
| Waist-Hip-Ratio (WHR) | 0.82 (0.76–0.89) | 0.82 (0.78–0.88) | 0.85 (0.80 – 0.91) | 0.85 (0.80 – 0.90) | 0.86 (0.81 – 0.91) | 0.86 (0.82 – 0.90) |
| Waist-Height-Ratio (WHtR) | 0.46 (0.42–0.53) | 0.45 (0.42–0.50) | 0.49 (0.46 – 0.54) | 0.44 (0.41 – 0.49) | 0.45 (0.42 – 0.48) | 0.45 (0.42 – 0.49) |
| Conicity Index (Cindex) | 1.21 (1.14–1.31) | 1.22 (1.15–1.30) | 1.22 (1.16 – 1.30) | 1.21 (1.14 – 1.28) | 1.22 (1.15 – 1.29) | 1.18 (1.12 – 1.25) |
| Body adiposity Index (BAI) | 26.9 (23.8 – 30.8) | 25.6 (23.3 – 28.3) | 27.5 (25.5 – 31.1) | 23.0 (19.9 – 25.3) | 21.7 (19.9 – 24.6) | 22.6 (20.7 – 25.6) |
| Body roundness index (BRI) | 2.70 (1.97–3.95) | 2.48 (1.93–3.36) | 3.19 (2.58–4.08) | 2.36 (1.84–3.25) | 2.40 (1.94–3.02) | 2.54 (2.03 – 3.28) |
| Body shape index (ABSI) | 0.81 (0.76 – 0.85) | 0.82 (0.77 – 0.86) | 0.79 (0.76 – 0.85) | 0.79 (0.75 – 0.85) | 0.81 (0.77 – 0.87) | 0.77 (0.73 – 0.82) |
| **Metabolic disorder, n (%)** |  |  |  |  |  |  |
| IFG/Diabetes | 42 (15.0) | 96 (15.2) | 58 (15.6) | 23 (16.6) | 42 (14.7) | 37 (17.5) |
| Hypertension | 117 (41.8) | 254 (40.2) | 152 (40.9) | 81 (58.3) | 152 (53.2) | 107 (50.5) |
| IFG/Diabetes/Hypertension | 133 (47.5) | 312 (49.4) | 183 (49.2) | 91 (65.5) | 170 (59.4) | 126 (59.4) |
| Metabolic Syndrome (MetS) | 127 (45.4) | 220 (34.8) | 196 (52.7) | 36 (25.9) | 60 (21.0) | 57 (26.9) |

**Table 2:** Correlation between the anthropometric measures.

|  |  | Male (n=139) |  |  |  |  |  |  |  |  |  |
| --- | --- | --- | --- | --- | --- | --- | --- | --- | --- | --- | --- |
|  |  | BW | WC | HC | BMI | WHR | WHtR | Cindex | BAI | BRI | ABSI |
| Female (n=280) | **Fulbe** |  |  |  |  |  |  |  |  |  |  |
|  | BW | ---- | 0.751* | 0.807* | 0.898* | 0.327* | 0.613* | 0.219* | 0.508* | 0.614* | -0.044 |
|  | WC | 0.748* | ----- | 0.741* | 0.744* | 0.716* | 0.941* | 0.755* | 0.595* | 0.942* | 0.523* |
|  | HC | 0.854* | 0.730* | ----- | 0.774* | 0.162 | 0.656* | 0.363* | 0.799* | 0.660* | 0.126 |
|  | BMI | 0.917* | 0.713* | 0.812* | ----- | 0.380* | 0.740* | 0.213* | 0.722* | 0.740* | -0.085 |
|  | WHR | 0.272* | 0.705* | 0.103 | 0.260* | ---- | 0.743* | 0.789* | 0.154 | 0.743* | 0.674* |
|  | WHtR | 0.653* | 0.952* | 0.677* | 0.736* | 0.689* | ---- | 0.754* | 0.705* | 0.998* | 0.520* |
|  | Cindex | 0.157* | 0.723* | 0.235* | 0.115* | 0.837* | 0.698* | ---- | 0.321* | 0.758* | 0.940* |
|  | BAI | 0.622* | 0.601* | 0.825* | 0.796* | 0.072 | 0.713* | 0.179* | ----- | 0.705* | 0.092 |
|  | BRI | 0.651* | 0.953* | 0.676* | 0.732* | 0.690* | 0.999* | 0.702* | 0.711* | ---- | 0.524* |
|  | ABSI | -0.176* | 0.423* | -0.073 | -0.244* | 0.711* | 0.390* | 0.917* | -0.125* | 0.394* | ---- |
|  |  | Male (n=286) |  |  |  |  |  |  |  |  |  |
| Female (n=632) | **Mbororo** |  |  |  |  |  |  |  |  |  |  |
|  | BW | ---- | 0.602* | 0.543* | 0.879* | 0.254* | 0.414* | -0.088 | 0.133* | 0.423* | -0.300* |
|  | WC | 0.657* | ---- | 0.530* | 0.550* | 0.698* | 0.891* | 0.666* | 0.288* | 0.912* | 0.478* |
|  | HC | 0.750* | 0.674* | ---- | 0.474* | -0.145* | 0.406* | 0.212* | 0.672* | 0.429* | 0.077 |
|  | BMI | 0.888* | 0.641* | 0.687* | ---- | 0.231* | 0.533* | -0.146* | 0.402* | 0.547* | -0.384* |
|  | WHR | 0.265* | 0.755* | 0.091* | 0.294* | ---- | 0.665* | 0.621* | -0.193* | 0.684* | 0.534* |
|  | WHtR | 0.525* | 0.943* | 0.591* | 0.653* | 0.754* | ---- | 0.676* | 0.471* | 0.977* | 0.446* |
|  | Cindex | 0.085* | 0.754* | 0.289* | 0.067 | 0.779* | 0.735* | ---- | 0.128* | 0.657* | 0.936* |
|  | BAI | 0.424* | 0.527* | 0.752* | 0.657* | 0.084* | 0.658* | 0.243* | ---- | 0.490* | 0.008 |
|  | BRI | 0.526* | 0.944* | 0.593* | 0.652* | 0.755* | 0.998* | 0.736* | 0.659* | ---- | 0.469* |
|  | ABSI | -0.197* | 0.504* | 0.054 | -0.244* | 0.649* | 0.485* | 0.935* | 0.022* | 0.487* | ---- |

|  |  | Male (n=212) |  |  |  |  |  |  |  |  |  |
| --- | --- | --- | --- | --- | --- | --- | --- | --- | --- | --- | --- |
|  | General population | BW | WC | HC | BMI | WHR | WHtR | Cindex | BAI | BRI | ABSI |
| Female (n=372) | BW | ---- | 0.573* | 0.698* | 0.887* | 0.007 | 0.405* | -0.082 | 0.391* | 0.404* | -0.332* |
|  | WC | 0.637* | ---- | 0.669* | 0.559* | 0.536* | 0.930* | 0.691* | 0.524* | 0.931* | 0.462* |
|  | HC | 0.794* | 0.663* | ----- | 0.652* | -0.165* | 0.573* | 0.218* | 0.742* | 0.573* | 0.003 |
|  | BMI | 0.899* | 0.663* | 0.717* | ----- | 0.038 | 0.533* | -0.099 | 0.614* | 0.534* | -0.374* |
|  | WHR | 0.063 | 0.617* | -0.095* | 0.108* | ----- | 0.559* | 0.696* | -0.064 | 0.562* | 0.632* |
|  | WHtR | 0.487* | 0.938* | 0.554* | 0.608* | 0.651* | ---- | 0.706* | 0.652* | 0.998* | 0.485* |
|  | Cindex | -0.072 | 0.656* | 0.128* | -0.077 | 0.761* | 0.675* | ---- | 0.232* | 0.707* | 0.950* |
|  | BAI | 0.493* | 0.549* | 0.775* | 0.679* | -0.045 | 0.653* | 0.157* | ----- | 0.653* | 0.034 |
|  | BRI | 0.486* | 0.939* | 0.554* | 0.607* | 0.652* | 0.998* | 0.677* | 0.652* | ---- | 0.486* |
|  | ABSI | -0.368* | 0.369* | -0.133* | -0.397* | 0.648* | 0.391* | 0.927* | -0.094 | 0.393* | ---- |

\*: statistically significant with p<0.05

**Table 2:**
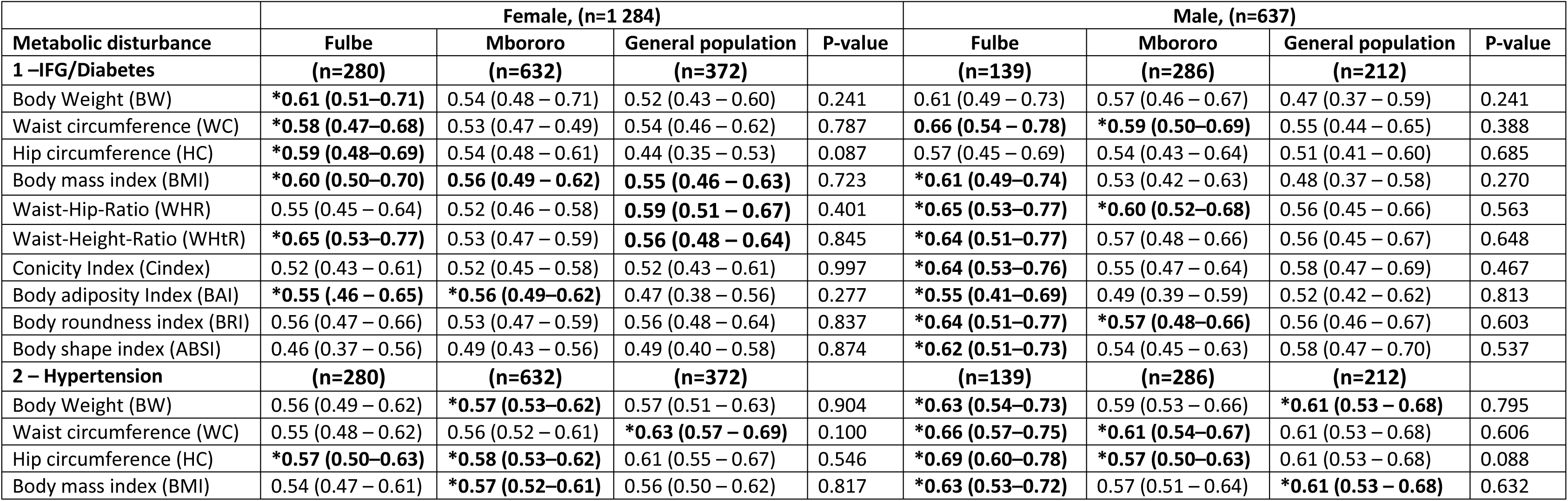
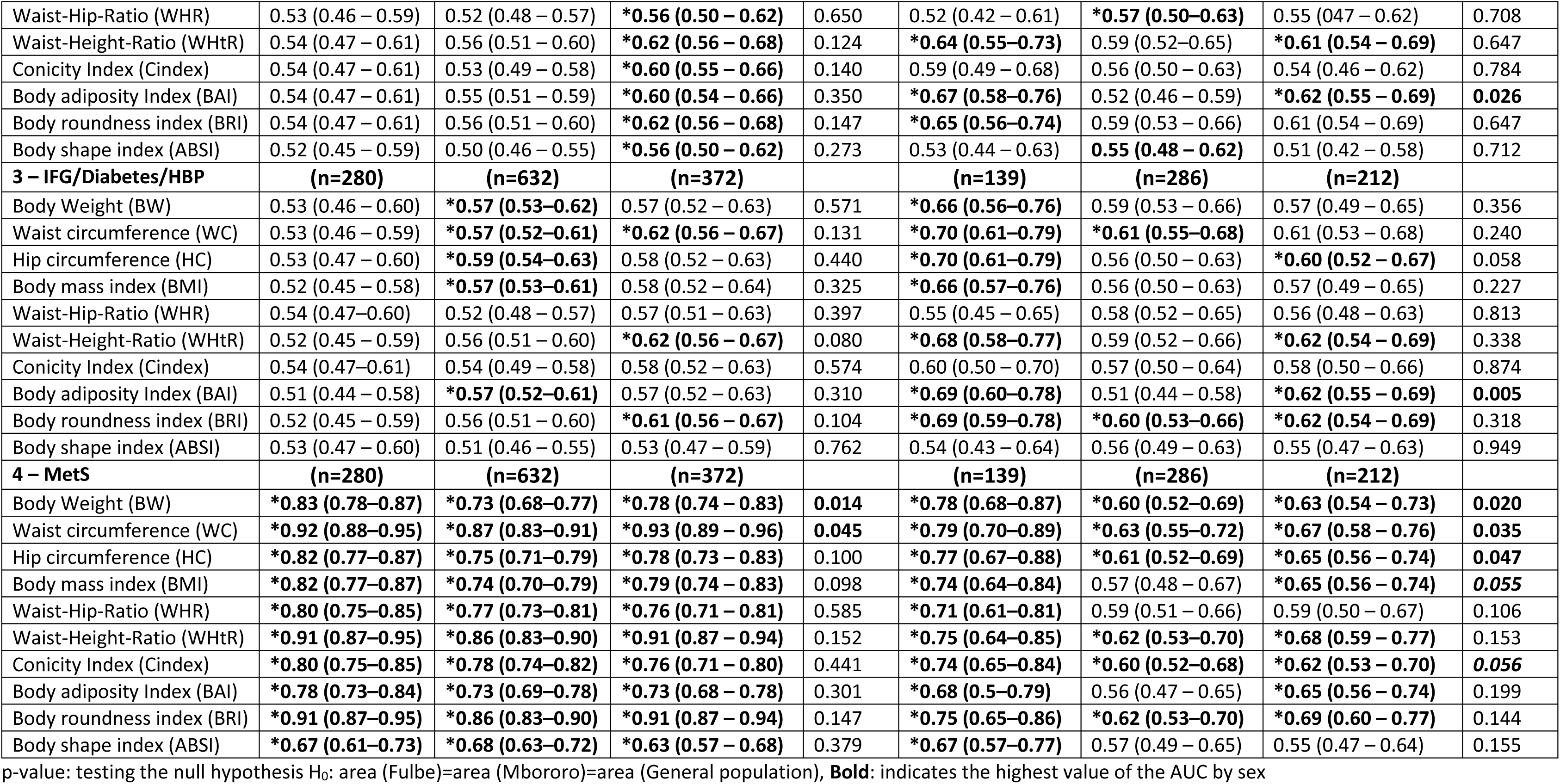
The area under the ROC curve (95%CI) of each anthropometric measurements for the IFG/diabetes, Hypertension, IFG/diabetes/hypertension and MetS.

**Table 3:**
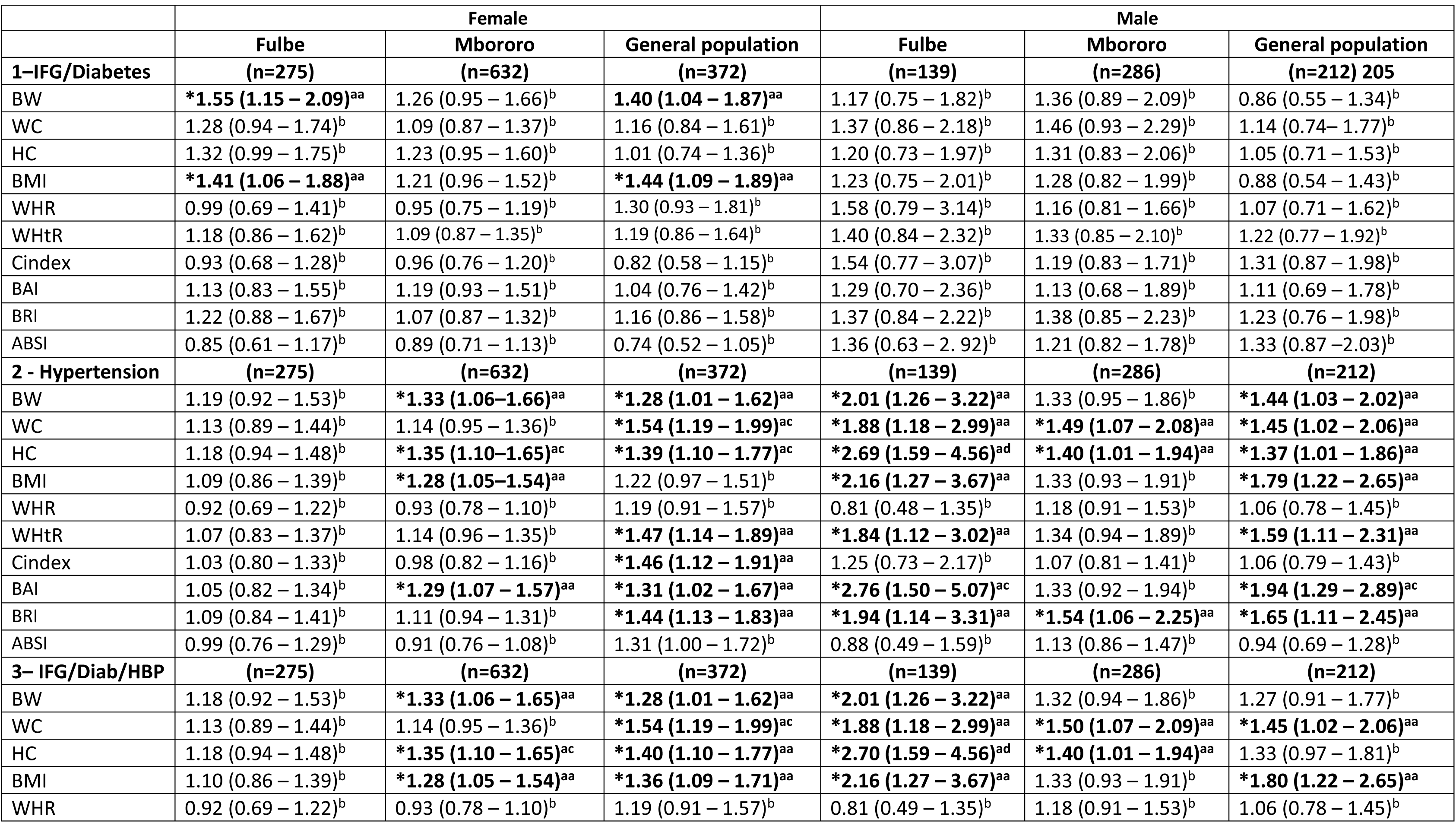
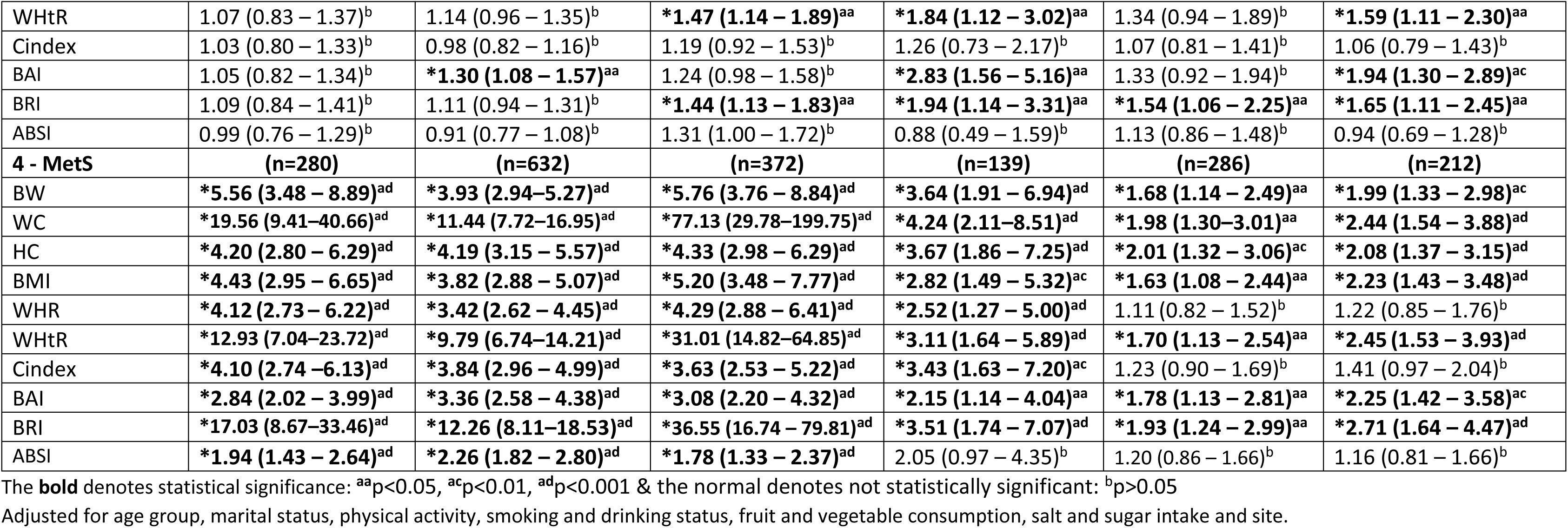
Multivariable Adjusted Odds Ratio (95% CI) for the presence of IFG/diabetes, Hypertension, IFG/diabetes/hypertension and MetS from multivariate logistic regression.

## Notes

### Competing Interest Statement

The authors have declared no competing interest.

### Funding Statement

This study was funded by World Diabetes Foundation.

### Author Declarations

The study protocol was approved by National Ethics Committee of Cameroon.

